# Prevalence, patterns, and socio-demographic risk factors of *Pf*HRP2 antigen among pregnant and non-pregnant populations in an urban setting: a retrospective study in Ghana

**DOI:** 10.1101/2025.08.21.25334142

**Authors:** Felix Osei-Boakye, Charles Nkansah, Samuel Kwasi Appiah, Gabriel Abbam, Charles Angnataa Derigubah, Emmanuel Ike Ugwuja, Boniface Nwofoke Ukwah, Victor Udoh Usanga, Theresa Ndago, Karikari Appau, Abdul-Razak Saasi, Ejike Felix Chukwurah

## Abstract

**Introduction:** Falciparum is the major contributor to malaria and causes the most lethal disease in humans globally. Although several government-funded interventions exist to help eliminate malaria, the disease persists and continues to afflict Ghanaians of all ages, including children, pregnant mothers, and adults.

**Methods:** This single-facility retrospective study was carried out at Sunyani Municipal Hospital. Patients’ malaria records spanning December 2020 and November 2021 were collected, analyzed, and visualized using IBM SPSS and GraphPad Prism. Proportions, associations, odds ratios, confidence intervals, and effect sizes were determined.

**Results:** Prevalence of malaria was 11.9%, with increased burden in males (14.5%), children 5-17 years (19.7%), pregnant women (17.2%), and in the rainy season (13.0%). Male sex (OR: 1.440, *p*<0.001), ages 5-17 years (OR: 2.538, *p*<0.001), <5 years (OR: 1.471, *p*<0.001), pregnancy (OR: 1.559, *p*=0.003), and rainy (OR: 1.251, *p*<0.001) were significant risk factors for falciparum malaria. Also, 78.2% (*p*<0.001) of cases were from the OPD irrespective of season (rainy [79.2%, *p*<0.045]; dry [76.8%, *p*<0.001]). Malaria was high in males in rainy (15.3%, *p*<0.001) and dry seasons (13.6%, *p*<0.001). Rainy (*r*=0.111) and dry seasons (*r*=0.115) showed a weak positive association with malaria.

**Conclusion:** *Pf*HRP2 antigen positivity was associated with sex, age, patient type, and season, with increased prevalence in males, children 5-17 years, pregnancy, and the rainy season. Age showed a weak positive association with *Pf*HRP2 antigen positivity, whereas sex, patient type, and season showed negligible effects. Male sex, children 5-17 years, <5 years, pregnancy, and rainy season were risk factors for falciparum malaria. Malaria was associated with the source of laboratory request, with the bulk of the malaria cases reported among OPD patients, regardless of season. Rainy and dry seasons showed a weak positive association with malaria when stratified by age.

## Introduction

Malaria is a deadly infectious disease caused by Plasmodium species, a protozoan parasite that is disseminated by a mosquito vector [1]. Out of the several species of Plasmodium that infect primates, *P. falciparum*, *P. malariae*, *P. ovale*, *P. vivax*, and *P. knowlesi* cause disease in humans [2]. Among these species, falciparum is the major contributor to malaria and causes the most lethal disease in humans globally [2]. Malaria is spread from person to person when a Plasmodium-infected female Anopheles mosquito, the vector, bites a susceptible human host [3, 4].

Malaria infects approximately 263 million people and kills 597,000 people worldwide annually [5]. These recent global estimates suggest 11 million more individuals were infected with malaria in 2023 than in the previous year [5]. The World Health Organization’s (WHO) designated African Region contributes the greatest proportion of the global malaria burden and deaths, with 94.0% and 95.0%, respectively, occurring in this region [5]. Furthermore, the WHO report for the year under review suggests a continuous rise in the estimate of new cases, with the WHO African Region reporting the highest increase for over two decades, followed by the Eastern Mediterranean Region (89.7% and 15.5%), respectively [5]. About 246 million people from the WHO African Region were infected with malaria, while 569,000 individuals suffered malaria-associated mortality in 2023 [5]. In the same year, about 34.0% out of 36 million pregnant women in this region were afflicted by malaria, whereas 73.7% of infected children below five years died [5]. Furthermore, in Ghana, Plasmodium impacts the health of children, causing over 20% of mortality and 40% of children to be admitted [6].

In conducive environments, the Plasmodium parasite can infect a single human host over a thousand times annually [7]. Due to the multiple exposures over time, adults rapidly gain immunity, but have a higher probability of succumbing to the disease than children [7]. Plasmodium infects humans of varied clinical characteristics and ages, with pregnant individuals and children below the age of 5 years being particularly more susceptible to malaria [8]. This non-discriminatory nature of the disease contributes to its life-threatening potential. The increased susceptibility of pregnant individuals is attributed in part to the increased secretion and release of certain substances, such as carbon dioxide, that attract the vector [9]. Whereas young children may have limited exposure to Plasmodium, and may therefore lack specific immunity and/or immune memory to the antigens/proteins of the parasite [10].

In sub-Saharan Africa, where malaria is common, pregnant individuals are challenged with enormous risks during pregnancy [11]. During this period, the Plasmodium-infected erythrocytes become confined in the placenta of the expectant mothers, and cause interference in the transfer of oxygen [4] and nutrients between the mother and fetus [4, 12], resulting in *in utero* retarded growth retardation of the fetus [12]. Other outcomes include abortions, preterm birth [4, 11, 13–15], stillbirth [4, 15], and low birth weight [11, 13, 14], anemia in the fetus [4] or mother [13], and death of the mother [14] and child [13]. In contrast, whereas children below the age of 5 years are likely to suffer from cerebral malaria that results in permanent intellectual growth impairment, older children are challenged with uncomplicated diseases [16]. The intraerythrocytic life cycle of the parasite causes a rapid increase in the parasite population in the bloodstream following the concomitant lysis of Plasmodium-infected erythrocytes [17, 18]. This malaria-induced hemolysis results in the rapid development of anemia [18, 19], a deadly complication of malaria that causes increased sickness and death among mothers and minors [20].

In support of the WHO’s vision for a malaria-free world, the Government of Ghana initiated programs to reduce the transmission of Plasmodium through several malaria preventive interventions, such as the provision of sulfadoxine-pyrimethamine prophylaxis for pregnant women, mass distribution of long-lasting insecticide-treated nets, and indoor residual spraying. Despite these interventions, malaria infection persists and continues to affect the population. Mohammed *et al.,* [21], using surveillance data spanning 2015 to 2019 in the same municipality, reported an overwhelmingly high malaria prevalence of 49.4%. Although the yearly trend data reported by Mohammed *et al.,* [21], suggest an increase in cases in months with heavy rainfall, the association, direction, and magnitude of the relationship remain unknown in the Bono Region and Ghana at large. Also, the study by Mohammed *et al.,* [21], failed to determine risk factors that favor malaria transmission, and the association between malaria and the source of laboratory requests. Furthermore, data on the burden of malaria in Sunyani are limited. Consequently, this study determined the prevalence of malaria and compared the burden across different vulnerable populations, identified sociodemographic risk factors, and determined their association with falciparum malaria in this setting. Furthermore, this study identified the risk factors, direction, and magnitude of the effect of these risk factors. The association between the source of the laboratory request and malaria was also explored.

## Participants and methods

### Study design and setting

This single-center retrospective cross-sectional study was hospital-based and conducted at the Sunyani Municipal Hospital between 29^th^ July and 14^th^ August 2025. The study retrospectively retrieved, analyzed, and visualized records of patients suspected of having malaria infection. All eligible patients’ records from 1^st^ December 2020 to 30^th^ November 2021 were included in the study. The Sunyani Municipal Hospital is the second largest government hospital in the city of Sunyani and the entire Sunyani municipality, with an inpatient capacity of 105 beds [22]. Services offered at the hospital include: obstetric and gynecologic, internal medicine, ophthalmic, pharmaceutical, medical diagnostics (clinical laboratory and imaging), and several other clinics, like the highly active antiretroviral therapy (HAART) center, infectious diseases, and antihypertensive and diabetic clinics. Sunyani is located in the Sunyani municipality and is the capital city of the Bono Region, one of sixteen regions in Ghana. The Sunyani municipality has a human population of about 123,224 inhabitants [22].

### Study population

The participants involved in the study were either symptomatic, asymptomatic, or pre-surgery patients who sought medical care at the outpatient units/clinics of the Sunyani Municipal Hospital or were admitted to any of its inpatient wards. All eligible records of such patients from 1^st^ December 2020 to 30^th^ November 2021 were accessed. The study population included children, non-pregnant adults, and pregnant women. A total of 12,598 records were retrieved from the laboratory’s archive, of which 4.62% (582) were excluded due to incomplete patient information. The resulting 12,016 records with complete patient information were considered eligible and were included in the study. The reasons for the exclusion of participant records were unavailability of either patients’ age, sex, source of laboratory request, test result for *Plasmodium falciparum* histidine-rich protein 2 (*Pf*HRP2) antigen, date test was performed, and/or indecipherable handwriting.

### Sample Size determination

The sample size of the population was computed using the StatCalc function in EpiInfo v7.2.2.2 (Centers for Disease Control and Prevention [CDC], United States). The statistical parameters used for the computation were: power of 80%, two-sided confidence level of 95%, ratio (unexposed: exposed) of 0.5, and a prevalence of 8.9% reported by Fondjo et al., [6] was used as the percentage outcome in the unexposed group, whereas 17.0% was assumed for the percentage outcome in the exposed group. The Fleiss w/CC value yielded the largest sample size of 680, but this was increased to 12,016 using convenient non-randomized sampling to include all eligible participants and to reduce bias.

### Data collection

Laboratory registers in which the results of patients’ routine malaria screening were entered were recovered from the facility’s archive, and the required data were extracted between 29^th^ July and 1^st^ August 2025. Patient records were collected using a non-randomized, convenience sampling method. The information was entered into a Microsoft Excel spreadsheet. Information retrieved from the records included date of screening, age, and sex of the patient, and the unit/ward from which the test was requested. The data was later screened to remove duplicate entries and records with missing data: records without either age, sex, source of request, test result for HRP2 antigen, or date of screening. The patients’ information collected included age, sex, source of laboratory request, test result for *Pf*HRP2 antigen, and the date the test was performed. However, the names and identification numbers that could enable patient identification were not collected. The screening for *Pf*HRP2 was performed using the SD BIOLINE *in vitro* immunodiagnostic point-of-care test (POCT) kit, with catalogue number: 05FK50 (Abbott Laboratories, United States).

### Ethical consideration and informed consent

The study was approved by the Committee on Human Research, Publication and Ethics (CHRPE) of the School of Medicine and Dentistry, Kwame Nkrumah University of Science and Technology (Reference: CHRPE/AP/689/25). Furthermore, permission was obtained from the medical director of Sunyani Municipal Hospital and the head of the laboratory department before commencing the study. However, due to the retrospective design employed, consent from the participants was not required.

### Statistical data analysis

The data were initially extracted onto a Microsoft Excel 2021 spreadsheet, which was later exported to IBM SPSS v27 (Armonk, NY: IBM Corp.). The age of participants in years was presented as median, with 25^th^ and 75^th^ percentiles following a significant (*p*≤0.001) Kolmogorov-Smirnov test for normality. Furthermore, age was recoded into four nominal categories. All nominal data, including sex, age (in years), patient type, and season, were reported as frequencies, with their percentages enclosed in parentheses. The Fisher’s Exact and Phi tests were used to determine the association and effect of *P. falciparum* HRP2 (*Pf*HRP2) antigen on sex, patient type, and season. Conversely, the Chi-square and Cramer’s V tests were used to determine the association between the *Pf*HRP2 antigen and age. The odds of *Pf*HRP2 antigen positivity and its respective confidence levels were determined using binary logistic regression. Bar charts were visualized using GraphPad Prism v8 (GraphPad Software, San Diego, California, USA). Statistical significance was set at *p*≤0.05 for all analyses.

## Results

### Socio-demographic characteristics of the participants

Table 1 shows the socio-demographic distribution of the participants. Out of the 12016 patients’ data retrieved, the majority (65.8% [7905]) were females, aged between 18 and 40 years (37.4% [4492]), non-pregnant (97.3% [11696]), and tested for malaria in the rainy season (51.4% [6172]). The median age of the patients was 23 years, with 25^th^ and 75^th^ percentiles of 6.0 and 39.0 years, respectively (Table 1).

**Table 1.**
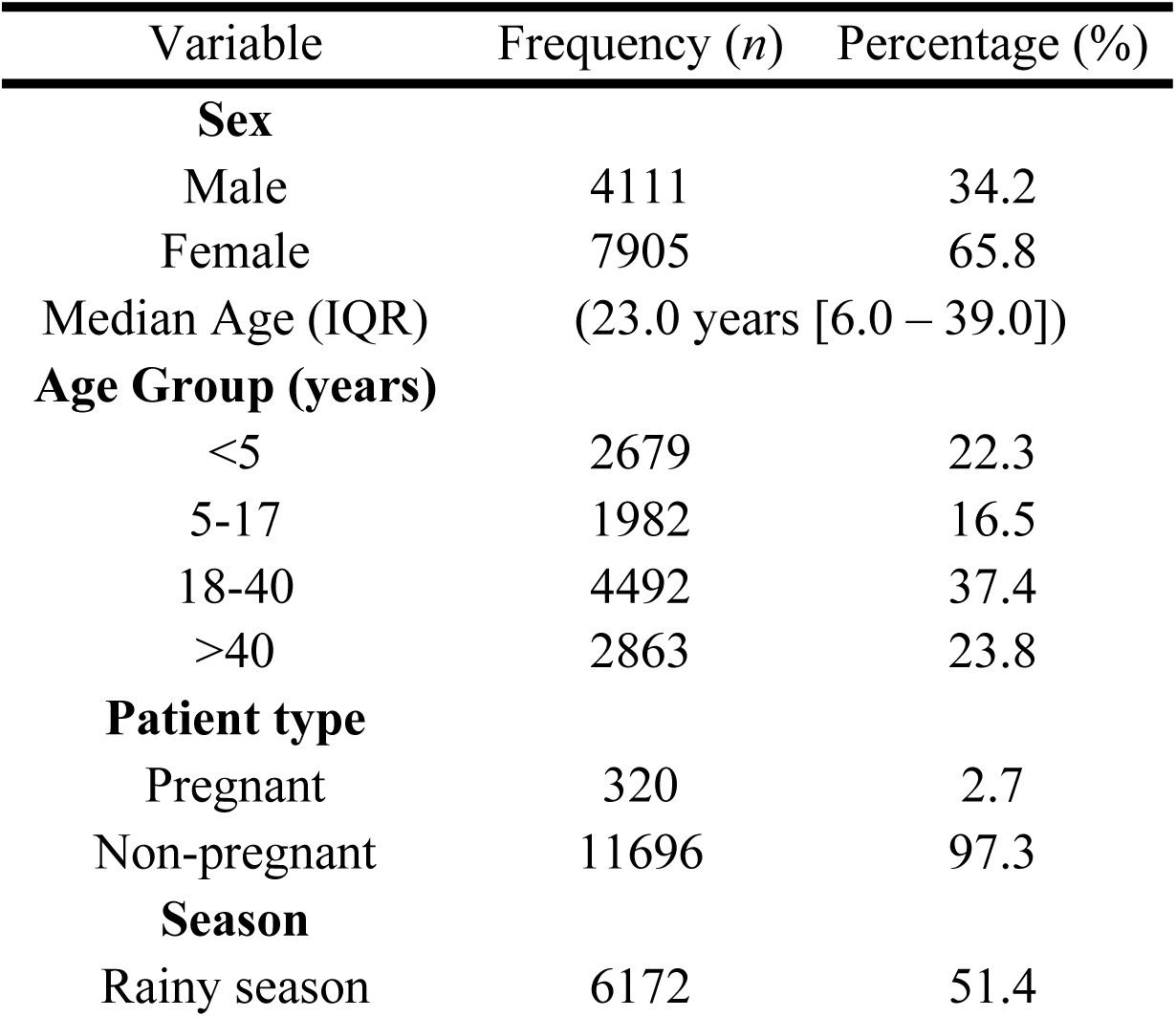
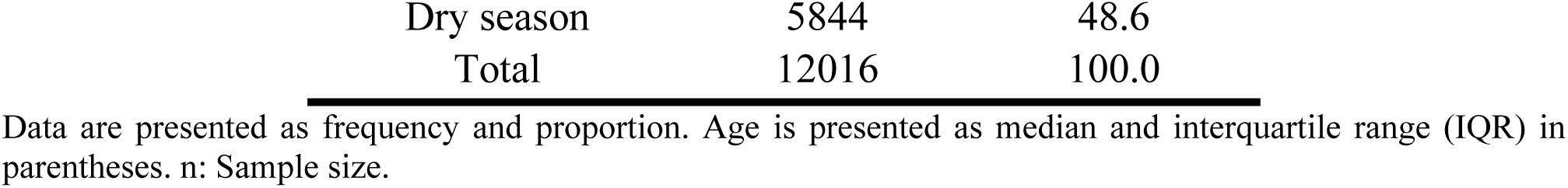
Socio-demographic characteristics of the participants at Sunyani Municipal Hospital, Ghana (December 2020-November 2021).

### Prevalence of *Pf*HRP2 antigen, and its association with socio-demographic factors

Of the 12016 suspected malaria cases, 11.9% (1429) were positive for *Pf*HRP2 antigen. Out of 4111 male patients suspected of having malaria, 14.5% (596) tested positive, whereas 10.5% out of the 833 females were positive. The prevalence of *Pf*HRP2 was increased (19.7% [390/1982]) among patients with ages 5-17 years, followed by <5 years (12.4% [333/2679]), 18-40 years (10.1% [454/4492]), and >40 years (8.8% [252/2863]). Furthermore, the prevalence of *Pf*HRP2 was increased (17.2% [55/320]) among pregnant populations, and (13.0% [804/6172]) in the rainy season. The *Pf*HRP2 positivity was significantly associated with sex (*p*<0.001), age (*p*<0.001), patient type (*p*=0.004), and season (*p*<0.001). Age had a significant and weak positive effect on *Pf*HRP2 (r=0.114, *p*<0.001), whereas sex, patient type, and season showed negligible effects on *Pf*HRP2 (Table 2).

**Table 2.** Prevalence of *Pf*HRP2 antigen, and its association with socio-demographic factors (December 2020-November 2021).

| Variable | Total<br>suspected<br>malaria<br>*n* (%) | Positive<br>*Pf*HRP2<br>*n* (%) | Negative<br>*Pf*HRP2<br>*n* (%) | *p* | Effect size |  |
| --- | --- | --- | --- | --- | --- | --- |
|  |  |  |  |  | Coefficient | *p* |
| **Sex** |  |  |  | **<0.001** | -0.058<sup>a</sup> | **<0.001** |
| Male | 4111 (34.2) | 596 (14.5) | 3515 (85.5) |  |  |  |
| Female | 7905 (65.8) | 833 (10.5) | 7072 (89.5) |  |  |  |
| **Age (years)** |  |  |  | **<0.001** | 0.114<sup>b</sup> | **<0.001** |
| <5 | 2679 (22.3) | 333 (12.4) | 2346 (87.6) |  |  |  |
| 5-17 | 1982 (16.5) | 390 (19.7) | 1592 (80.3) |  |  |  |
| 18-40 | 4492 (37.4) | 454 (10.1) | 4038 (89.9) |  |  |  |
| >40 | 2863 (23.8) | 252 (8.8) | 2611 (91.2) |  |  |  |
| **Patient type** |  |  |  | **0.004** | -0.027<sup>a</sup> | **0.003** |
| Pregnant | 320 (2.7) | 55 (17.2) | 265 (82.8) |  |  |  |
| Non-pregnant | 11696 (97.3) | 1374 (11.7) | 10322 (88.3) |  |  |  |
| **Season** |  |  |  | **<0.001** | -0.036<sup>a</sup> | **<0.001** |
| Rainy | 6172 (51.4) | 804 (13.0) | 5368 (87.0) |  |  |  |
| Dry | 5844 (48.6) | 625 (10.7) | 5219 (89.3) |  |  |  |
| **Total** | 12016 (100.0) | 1429 (11.9) | 10587 (88.1) |  |  |  |

Data are presented as frequencies with corresponding proportions in parentheses. *Pf*HRP2: *Plasmodium falciparum* Histidine-rich protein 2; *p*: P-value; <sup>a</sup> Phi; <sup>b</sup> Cramer's V; *p* was significant at $\leq 0.05$ .

### Socio-demographic risk factors of *Pf*HRP2 antigen among patients at Sunyani Municipal Hospital, Ghana

Table 3 presents the socio-demographic risk factors for falciparum malaria among malaria-suspected patients. Male patients were 1.440 times (*p*<0.001) more vulnerable to falciparum infection than females. Patients aged 5-17 years and <5 years were 2.538 times (*p*<0.001) and 1.471 times (*p*<0.001) more susceptible to malaria infection, respectively. Whereas participants were 1.251 times (*p*<0.001) more susceptible to malaria in the rainy season, the pregnant population had 1.559 times (*p*=0.003) increased risk of falciparum infection than the non-pregnant group (Table 3).

**Table 3.** Socio-demographic risk factors of *Pf*HRP2 antigen among patients at Sunyani Municipal Hospital, Ghana (December 2020-November 2021).

| Variable | Binary logistic regression |  |
| --- | --- | --- |
|  | OR (95% CI) | *p* |
| **Sex** |  |  |
| Male | 1.440 (1.286-1.611) | **<0.001** |
| Female | 1.0 (reference) |  |
| **Age (years)** |  |  |
| $<5$ | 1.471 (1.237 – 1.748) | **<0.001** |
| 5-17 | 2.538 (2.141 - 3.009) | **<0.001** |
| 18-40 | 1.165 (0.991 - 1.369) | 0.064 |
| $>40$ | 1.0 (reference) | |
| **Patient type** |  |  |
| Pregnant | 1.559 (1.160 - 2.096) | **0.003** |
| Non-pregnant | 1.0 (reference) |  |
| **Season** |  |  |
| Rainy | 1.251 (1.119 - 1.398) | **<0.001** |
| Dry | 1.0 (reference) |  |

The data are presented as Odds ratio (OR), with 95% confidence interval (CI) in parentheses. *Pf*HRP2: *Plasmodium falciparum* histidine-rich protein 2; *p* was significant at $\leq 0.05$ .

### The association between *Pf*HRP2 antigen positivity and the source of the laboratory request

Table 4 presents the association between the source of laboratory request and malaria, stratified by season. Out of the 12016 patients included in the study, Outpatient Department (OPD) attendance accounted for the majority, 81.3% (9775). Furthermore, out of the total 1429 positive malaria cases, the majority (78.2% [1117]) was recorded among outpatients, followed by accident and emergency ward (15.3% [219]), maternity ward (2.6% [37]), pediatric ward (1.3% [19]), antenatal clinic (1.3% [18]), females’ ward (1.0% [14]), males’ ward (0.3% [4]), and post-natal clinic (0.1% [1]). There was a statistically significant association between malaria burden and the source of laboratory request (*p*<0.001). An increased proportion of the malaria cases recorded in both the rainy and dry seasons were from the OPD, followed by emergency and maternity. However, whereas in the rainy season, this was followed by antenatal, females’, pediatric, and males’ wards; the maternity ward was followed by pediatric, antenatal, and both females’ ward and post-natal clinic in the dry season. Furthermore, both the dry and rainy seasons showed negligible effects, although significant ([*r*=0.068, *p*<0.001] and [*r*=0.051, *p*=0.045], respectively) on malaria across the different units/clinics (Table 4).

**Table 4.**
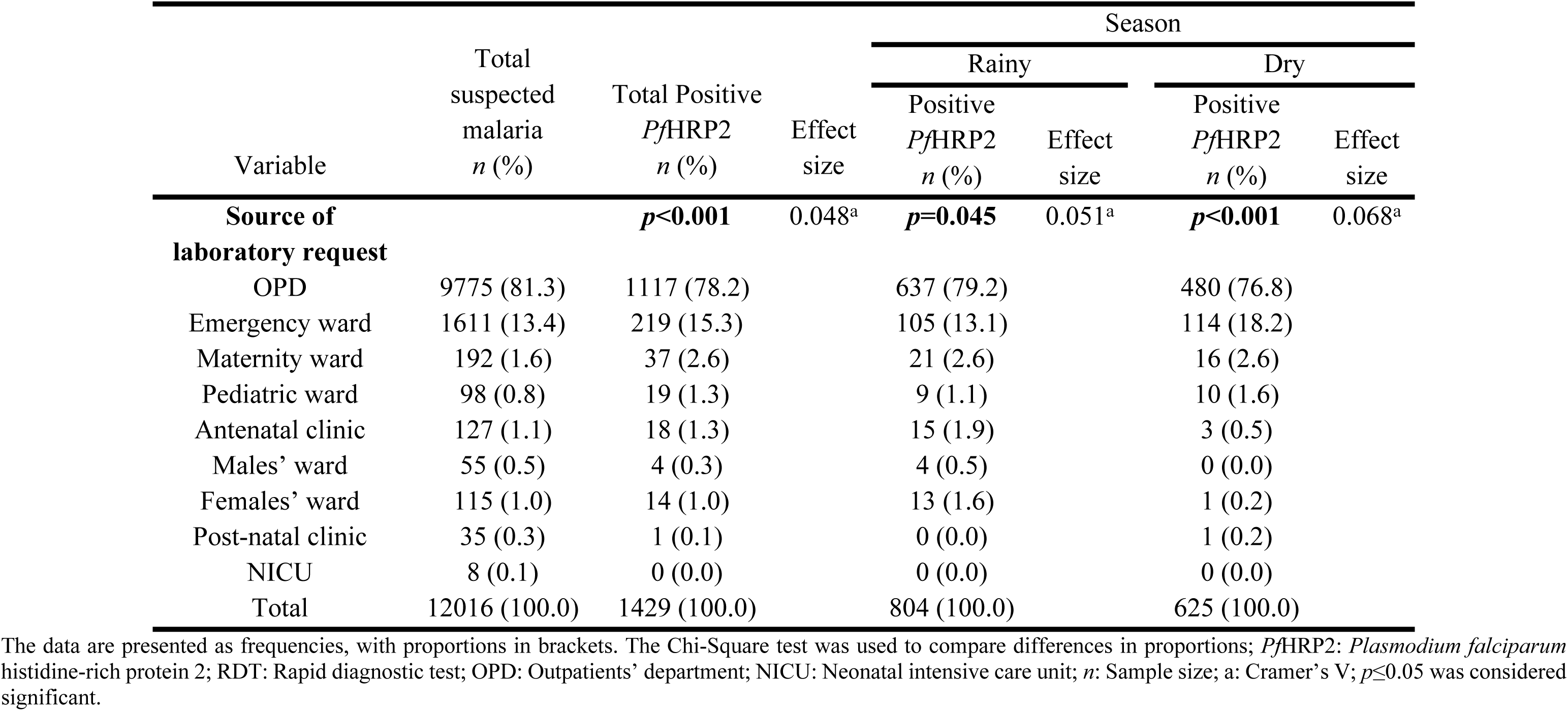
Association between source of laboratory request and *Pf*HRP2 antigen, stratified by season (December 2020-November 2021).

### Qualitative patterns of *Pf*HRP2 antigen across socio-demographic factors

The prevalence of malaria was significantly increased among males than females in both the rainy (15.3% [333/2175] vs 11.8% [471/2175]) and dry seasons (13.6% [263/1936] vs 9.3% [362/3908]). Increased percentage (21.4% [36/168] vs 12.8% [768/6004], respectively) of pregnant than non-pregnant women tested positive for PfHRP2 antigen in the rainy season. Similarly, increased percentage (12.5% [19/152] vs 10.6% [606/5692]) of pregnant than non-pregnant women tested positive for *Pf*HRP2 antigen, although not significant. Increased percentage (20.7% [224/1081]) of patients aged between 5-17 years tested positive for HRP2 antigen in the rainy season, followed by <5 years (13.6% [184/1357]), 18-40 years (10.8% [251/2328]), and > 40 years (10.3% [145/1406]). However, in the dry season, 18.4% (166/901) of patients between 5-17 years were positive for HRP2, followed by <5 years (11.3% [149/1322]), 18-40 years (9.4% [203/2164]), and >40 years (7.3% [107/1457]). When stratified by sex, *Pf*HRP2 antigen positivity showed significant associations with both rainy (*p*<0.001) and dry seasons (*p*<0.001). Also, when stratified by age, *Pf*HRP2 antigen positivity showed significant associations with rainy (*p*<0.001) and dry seasons (*p*<0.001). However, PfHRP2 antigen showed significant association with rainy season (*p=*0.002), but not dry season. Furthermore, both rainy and dry seasons showed weak positive associations with PfHRP2 antigen positivity (*r*=0.111 and *r*=0.115, respectively) (Fig 1).

**Fig 1.**
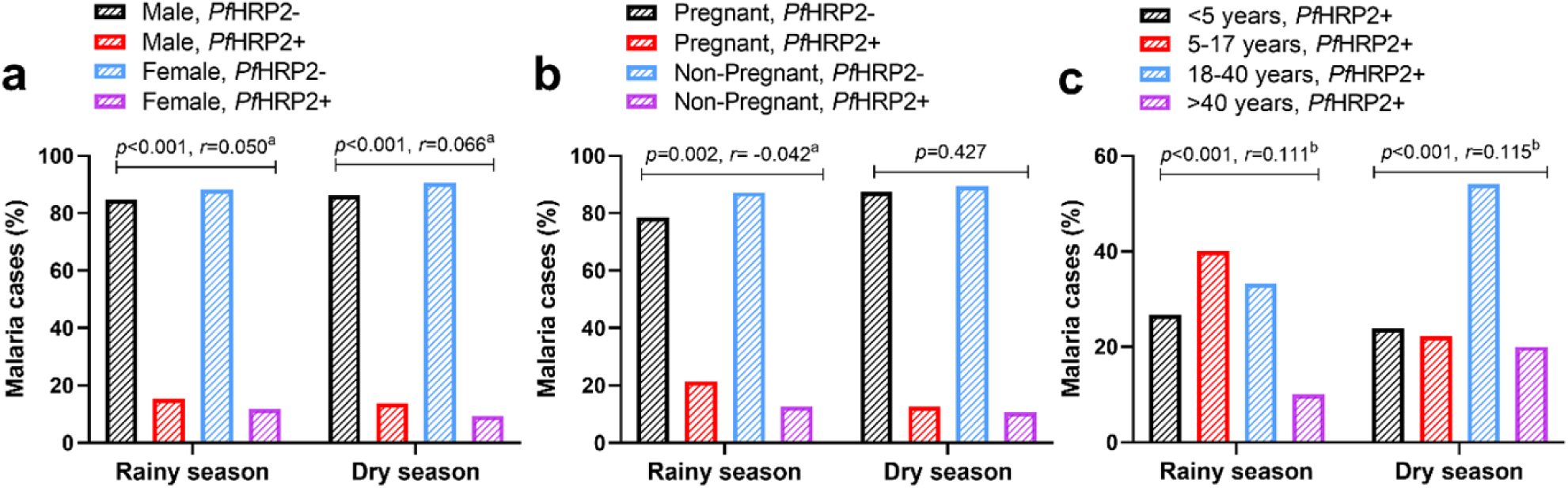
Qualitative patterns of *Pf*HRP2 antigen across socio-demographic factors, stratified by season. *Pf*HRP2: *Plasmodium falciparum* histidine-rich protein 2; +: Positive test; −: Negative test; a: Phi coefficient; b: Cramer’s V coefficient; *p* was significant at ≤0.05.

## Discussion

This study determined the prevalence, patterns, and socio-demographic risk factors of falciparum malaria among hospitalized and non-hospitalized patients using a rapid screening immunoassay. The prevalence of falciparum malaria by *Pf*HRP2 antigen positivity in this study was 11.9%, which is similar to 12.0% reported by Kwofie et al., [23] in Ghana’s capital city, Accra using a similar method. The similarity of the burden of malaria in the present study to that of Kwofie et al., [23] suggests that the burden of malaria in Sunyani was high. This is because the participants involved in this study were both symptomatic and asymptomatic patients who may share similar exposures as the subjects in the study by Kwofie et al., [23]. It is worth noting that the study population recruited by Kwofie et al., [23] were head porters who are vulnerable and marginalized people in society who are exposed to harsh environmental conditions. For instance, due to rural-urban migration, such individuals become destitute in major cities without proper accommodation, and are therefore exposed to plasmodial vector-breeding conditions.

The *Pf*HRP2 antigen positivity was significantly associated with sex, with a significantly increased prevalence in males (*p*<0.001). Furthermore, males were (OR: 1.440, *p*<0.001) more susceptible to falciparum malaria, although more females than males were suspected of malaria before testing. Briggs et al., [24] posit that biological differences associated with sex account in part for the varied immune response to plasmodial parasites and other pathogens. The plausible explanation for these immunological differences is related to the X chromosome; a greater number of immunoregulatory genes are found on the X chromosome [25], which is highly expressed in females [24]. This causes the immunologic reactions of the X chromosome to be skewed in favor of females [25]. In addition, hormonal differences between the sexes could alter immunological responses to pathogens in humans [25], whereas behavioral variables have been implicated in the male susceptibility to malaria [24]. In contrast to our findings, Boadu et al., [26] rather, report increased burden of malaria in females and attribute this to the peculiar fashion trend among females, in which females dress to reveal some parts of their bodies as they go about their routine tasks outside.

There was a significant weak positive association between age and *Pf*HRP2 antigen positivity. Furthermore, older children between ages 5-17 years were more afflicted, and were 2.538 times (*p*<0.001) more susceptible to falciparum malaria than children <5 years of age and adults. This corroborates the findings of Walldorf et al., [27], who reported increased risk of malaria in a similar population in Malawi. This finding further buttresses the malaria prevalence reported by Mensah et al., [28] among school-going children of similar ages in two different Ghanaian ecological settings: Begoro in the Eastern Region and Cape Coast in the Central Region. Children in this age range are usually of school-going age, and are more likely to be exposed to vector breeding environments as they stay and play outdoors. In addition, Walldorf et al., [27] suggest that older children seldom adhere to malaria preventive measures, such as not sleeping in insecticide-treated nets, causing them to be increasingly exposed to mosquito bites while indoors. Furthermore, the ongoing administration of prophylactic malaria vaccine to young children of ages 5 months to 3 years in Ghana since 2023 [29], offers protection against malaria and makes them less susceptible than older children who are not immunized against malaria.

The type of patient (pregnant or non-pregnant) was significantly associated (*p*=0.004) with *Pf*HRP2 antigen positivity, with increased positivity among the pregnant population. These findings corroborate the findings of Dwumfour et al., [30] reported in Ejisu, Ghana, among a similar population. Also, pregnant women were 1.559 times (*p*=0.003) more susceptible to falciparum malaria compared to non-pregnant women. Pregnant women are increasingly susceptible to malaria [31], because they release chemoattractants that increase their attractiveness to the mosquito vector than non-pregnant women [32]. The ramification of this are increased mosquito bites that facilitate the transmission of Plasmodium species in pregnant women.

In this study, differences in season were significantly associated with falciparum malaria, with increased prevalence in the rainy season, and is consistent with the findings of Dicko et al., [33], whose study suggests an increased prevalence of malaria-induced fever in the rainy season than the dry season. Furthermore, the current study revealed a 1.251 times increased susceptibility to malaria in the rainy season than the dry season (*p*<0.001). The occurrence and transmission of several diseases in humans and other primates are affected by climate dynamics [34]. Fall *et al.,* [34], further posit that since malaria is a vector-borne disease, variables of climate change, such as rainfall and temperature can influence environmental conditions like vector breeding sites, which impacts the burden of malaria by altering the developmental life cycle of both the mosquito vector and Plasmodium (the parasite) within the vector [34]. Rainfall creates suitable breeding sites [26, 34, 35] to support the development of the aquatic forms of mosquitoes that eventually transform into blood-feeding adult mosquitoes, capable of spreading sporozoites of Plasmodium species.

The prevalence of malaria was significantly higher among patients who reported to the OPD than in the inpatient wards and other clinics. This corroborates the findings of Minwuyelet and Aschale [37] that reports a higher malaria burden among OPD patients in Ethiopia. And, although this finding was not explained, the bulk of the patients first report to the OPD before they are either admitted to the wards or referred to other clinics for management following a diagnosis. Furthermore, children playing in the rain or outdoors after raining is a common observation in Ghana, this may expose children of school-going age to vector-breeding sites and may have contributed to the increased malaria prevalence in children 5-17 years. However, in the dry season, many young adults aged 18-40 tend to spend much time outdoors for social gatherings. A ramification of this is exposure to increased mosquito bites, which impacts the susceptibility of this population to malaria. Conversely, the reason for the increased susceptibility of pregnant women and males during either rainy or dry seasons is not understood and requires further research.

Despite the relevance of this study, it had some limitations that future research could address. Firstly, the *Pf*HRP2 antigen rapid diagnostic test is specific to only *Plasmodium falciparum*; therefore, the presence of other human Plasmodium species may have been missed. Consequently, the burden of malaria in this setting may have been underestimated. Secondly, the study did not capture whether some of the patients had received malaria preventive interventions. For instance, information was lacking on whether the pregnant women were attending the antenatal clinic for the first time (registrant), or were repeat patients who had received sulfadoxine-pyrimethamine prophylaxis during previous visits. Thirdly, Fernández-Grandon et al., [9], posit that an increase in body mass can provide a large surface area that facilitates attraction of the vector to the human host. However, information on patients’ anthropometrics was lacking due to the retrospective nature of the study. Finally, the qualitative assay could not quantify the actual degree of the parasitemia. Therefore, we implore future studies to help bridge these gaps in literature.

## Conclusion

The *Pf*HRP2 antigen positivity was significantly associated with sex, age, patient type, and season, with increased prevalence in males, children 5-17 years, pregnant women, and rainy season. Age showed a weak positive association with *Pf*HRP2 antigen positivity, whereas, sex, patient type, and season showed negligible effects. Male sex, children 5-17 years, under 5 years, pregnancy, and rainy season were significant risk factors for falciparum malaria., Malaria was significantly associated with the source of laboratory request, with the majority of the malaria cases reported among OPD patients, regardless of season. Whereas more children 5-17 years were infected in the rainy season, more adults 18-40 years were infected with malaria in the dry season. Rainy and dry seasons showed weak positive associations with malaria when stratified by age.

## Data Availability

The dataset supporting this study has been deposited in the Zenodo online repository and available at https://doi.org/10.5281/zenodo.16887880.

## Acknowledgements

The authors wish to appreciate Dr. Robert Arkoh (Medical Superintendent), Mr. Ofori Agyei (Health Services Administrator), and all the Laboratory staff of Sunyani Municipal Hospital.

## Authors’ Contributions

**Conceptualization:** Felix Osei-Boakye, Charles Nkansah, Samuel Kwasi Appiah

**Data curation:** Theresa Ndago, Karikari Appau, Charles Angnataa Derigubah, and Abdul-Razak Saasi

**Formal analysis:** Felix Osei-Boakye

**Methodology:** Felix Osei-Boakye, Gabriel Abbam, Charles Angnataa Derigubah, Abdul-Razak Saasi

**Resources:** Felix Osei-Boakye

**Supervision:** Ejike Felix Chukwurah, Emmanuel Ike Ugwuja

**Validation:** Felix Osei-Boakye

**Visualization:** Felix Osei-Boakye

**Writing – original draft:** Felix Osei-Boakye, Charles Nkansah, Gabriel Abbam, Samuel Kwasi Appiah, Boniface Nwofoke Ukwah, Victor Udoh Usanga, Emmanuel Ike Ugwuja, and Ejike Felix Chukwurah

**Writing – review & editing:** Felix Osei-Boakye, Charles Nkansah, Gabriel Abbam, Samuel Kwasi Appiah, Boniface Nwofoke Ukwah, Victor Udoh Usanga, Abdul-Razak Saasi, Emmanuel Ike Ugwuja, and Ejike Felix Chukwurah, Charles Angnataa Derigubah, Theresa Ndago, and Karikari Appau

## Supporting information

S1 Data. Dataset used for the study.

